# Transforming Obesity Prevention for CHILDren (TOPCHILD) Collaboration: protocol for a systematic review with individual participant data meta-analysis of behavioural interventions for the prevention of early childhood obesity

**DOI:** 10.1101/2020.12.17.20248441

**Authors:** Kylie E Hunter, Brittany J Johnson, Lisa Askie, Rebecca K Golley, Louise A Baur, Ian C Marschner, Rachael W Taylor, Luke Wolfenden, Charles T Wood, Seema Mihrshahi, Alison J Hayes, Chris Rissel, Kristy P Robledo, Denise A O’Connor, David Espinoza, Lukas P Staub, Paul Chadwick, Sarah Taki, Angie Barba, Sol Libesman, Mason Aberoumand, Wendy A Smith, Michelle Sue-See, Kylie D Hesketh, Jessica L Thomson, Maria Bryant, Ian M Paul, Vera Verbestel, Cathleen Odar Stough, Li Ming Wen, Junilla K Larsen, Sharleen L O’Reilly, Heather M Wasser, Jennifer S Savage, Ken K Ong, Sarah-Jeanne Salvy, Mary Jo Messito, Rachel S Gross, Levie T Karssen, Finn E Rasmussen, Karen Campbell, Ana Maria Linares, Nina Cecilie Øverby, Cristina Palacios, Kaumudi J Joshipura, Carolina González Acero, Rajalakshmi Lakshman, Amanda L Thompson, Claudio Maffeis, Emily Oken, Ata Ghaderi, Maribel Campos Rivera, Ana B Perez-Exposito, Jinan C Banna, Kayla de la Haye, Michael Goran, Margrethe Røed, Stephanie Anzman-Frasca, Anna Lene Seidler, on behalf of the Transforming Obesity Prevention for CHILDren (TOPCHILD) Collaboration

## Abstract

**Introduction:** Behavioural interventions in early life appear to show some effect in reducing childhood overweight and obesity. However, uncertainty remains regarding their overall effectiveness, and whether effectiveness differs among key subgroups. These evidence gaps have prompted an increase in very early childhood obesity prevention trials worldwide. Combining the individual participant data (IPD) from these trials will enhance statistical power to determine overall effectiveness and enable examination of intervention-covariate interactions. We present a protocol for a systematic review with IPD meta-analysis to evaluate the effectiveness of obesity prevention interventions commencing antenatally or in the first year after birth, and to explore whether there are differential effects among key subgroups.

**Methods and analysis:** Systematic searches of Medline, Embase, CENTRAL, CINAHL, PsycInfo, and trial registries for all ongoing and completed randomised controlled trials evaluating behavioural interventions for the prevention of early childhood obesity have been completed up to March 2020 and will be updated annually to include additional trials. Eligible trialists will be asked to share their IPD; if unavailable, aggregate data will be used where possible. An IPD meta-analysis and a nested prospective meta-analysis (PMA) will be performed using methodologies recommended by the Cochrane Collaboration. The primary outcome will be body mass index (BMI) z-score at age 24 +/- 6 months using World Health Organisation Growth Standards, and effect differences will be explored among pre-specified individual and trial-level subgroups. Secondary outcomes include other child weight-related measures, infant feeding, dietary intake, physical activity, sedentary behaviours, sleep, parenting measures and adverse events.

**Ethics and dissemination:** Approved by The University of Sydney Human Research Ethics Committee (2020/273) and Flinders University Social and Behavioural Research Ethics Committee (project no. HREC CIA2133-1). Results will be relevant to clinicians, child health services, researchers, policy-makers and families, and will be disseminated via publications, presentations, and media releases.

**Registration:** Prospectively registered on PROSPERO: CRD42020177408

**STRENGTHS AND LIMITATIONS OF THIS STUDY:**

- This will be the largest individual participant data (IPD) meta-analysis evaluating behavioural interventions for the prevention of early childhood obesity to date, and will provide the most reliable and precise estimates of early intervention effects to inform future decision-making.
- IPD meta-analysis methodology will enable unprecedented exploration of important individual and trial-level characteristics that may be associated with childhood obesity or that may be effect modifiers.
- The proposed innovative methodologies are feasible and have been successfully piloted by members of our group.
- It may not be possible to obtain IPD from all eligible trials; in this instance, aggregate data will be used where available, and sensitivity analyses will be conducted to assess inclusion bias.
- Outcome measures may be collected and reported differently across included trials, potentially increasing imprecision; however, we will harmonise available data where possible, and encourage those planning or conducting ongoing trials to collect common core outcomes following prospective meta-analysis methodology.

## INTRODUCTION

Childhood obesity is one of the most serious public health issues of the 21^st^ century, and requires urgent action.^1, 2^ Globally, an estimated 38 million (6%) children aged under five years were living with overweight or obesity in 2019,^3^ and prevalence is increasing across every continent as environments become more obesity conducive.^4, 5^ While childhood obesity affects all sections of society, it disproportionately affects racial and ethnic minority groups^6, 7^ and populations with a lower socioeconomic position (SEP), and thus is also a major health equity issue.^4^ Children with obesity are much more likely to have obesity across the lifecourse,^8, 9^ and are at increased risk of short and long term negative health sequelae, such as poor mental and musculoskeletal health, type 2 diabetes, asthma, and cardiovascular disease.^10, 11^ This places a large burden on healthcare systems,^12^ and has significant economic consequences arising from increased disability and decreased productivity and life expectancy.^13^ Thus, identifying modifiable behaviours for the early prevention of childhood obesity is critical to inform the development of early intervention strategies.

There are a variety of modifiable behaviours that may influence energy balance and therefore may be implicated in childhood obesity prevention, namely, feeding practices, dietary intake, physical activity, sedentary behaviours and sleep. For instance, appropriate responsive feeding has been identified as promising for obesity prevention,^14-16^ while consumption of sugar sweetened beverages is associated with severe obesity in children aged less than 5 years.^17^ Data are mixed on the protective benefits of breastfeeding for the prevention of obesity, though some studies suggest that longer duration of exclusive breastfeeding may provide modest protection.^18-23^ Similarly, there may be an association between age at introduction of solids and growth,^24^ with mixed results surrounding the direction of this association and the underlying causal mechanisms. Previous systematic reviews have reported significant inverse associations between physical activity and measures of adiposity in children.^25-27^ Conversely, sedentary behaviours such as television viewing or screen time are associated with higher body mass index (BMI) levels^28, 29^ and greater adiposity^30^ in young children. There is now also a large body of observational evidence supporting the relationship between short sleep duration and an increased risk of obesity across all age groups, including infants and young children,^31-35^ though a recent systematic review found inconsistent evidence of an association between longer infant sleep duration and healthier body composition up to age 24 months.^36^

In addition to these behaviours, individual-level covariates known or hypothesised to be predictive for childhood obesity include pre-pregnancy maternal and paternal BMI, age, race, ethnicity, socioeconomic position, excess gestational weight gain, parity, smoking during pregnancy, gestational diabetes, birth mode of delivery (caesarean, vaginal), birthweight, gestational age at birth, baby’s sex, and childcare attendance. ^6, 7, 22, 37^ Some of these covariates may also be individual-level effect modifiers, prediciting how effective an intervention is likely to be, e.g. socioeconomic position and race/ethnicity. Trial-level characteristics such as timing of intervention onset, setting and the level of well-child health care available in the community may also modify intervention effectiveness.^38^

### Limitations and evidence gaps identified in previous reviews

In the past five years there have been numerous reviews of childhood obesity prevention trials encompassing a variety of intervention types, settings and age groups.^14, 39-45^ Few of these focused solely on infancy, and many spanned multiple life stages from the prenatal period to 18 years of age. One review found that family-based childhood obesity prevention interventions most frequently targeted children 2-10 years of age (78%), with fewer targeting infants aged 0-1 year (24%) or the prenatal period (8%).^39^ Most reviews highlighted the urgent need for further rigorous evidence to inform obesity prevention interventions in the very early childhood years.^14, 39-43, 45^ Given the consequences of rapid early life weight gain, associated epigenetic changes and early onset of obesity in many children,^3, 46, 47^ there is strong rationale to start preventive interventions early when biology is most amenable to change, and before negative obesity-conducive behavioural patterns are established.^2^

Most of the childhood obesity prevention reviews to date have used qualitative methodology such as narrative reviews, content analysis, and systematic reviews without meta-analysis to describe variations in study design, setting, population, interventions, and outcomes, and to hypothesise that certain individual and trial-level characteristics may enhance effectiveness via proposed conceptual frameworks and intervention models.^14, 39, 41-45^ Yet, quantitative evaluation is required to formally test these hypotheses. Recently, Brown et al^40^ updated a Cochrane systematic review and aggregate data meta-analysis on obesity prevention in children aged 0-18 years, and found that interventions focusing on diet and physical activity combined can lead to a small reduction in BMI z-score in children aged 0-5 years of age (mean difference -0.07, 95% CI -0.14 to -0.01). However, a huge variety in intervention approaches limited their ability to conduct meaningful comparisons, and many multicomponent interventions were originally reported as a whole package, precluding evaluation of discrete intervention characteristics. Moreover, the aggregated data were insufficient to derive conclusions on effect differences by individual-level characteristics such as ethnicity and socioeconomic position.

The Early Prevention of Obesity in Children (EPOCH) Collaboration conducted a world-first IPD prospective meta-analysis (PMA) of four randomised controlled trials (RCTs) of behavioural interventions for the prevention of early childhood obesity.^38^ They found that, compared with usual care, early childhood interventions were modestly effective in reducing BMI z-score 18-24 months after birth by 0.12 standard deviations (which translates to a 2% decrease in obesity prevalence). However, when accounting for missing data this difference was no longer significant. There was some heterogeneity across trials, and interventions appeared to be more effective in populations with limited publicly funded existing health care programs, in this instance defined as a maximum of one postnatal home visit.^38^ However, this finding needs to be confirmed in analyses including more than four studies. EPOCH’s predictive analyses of individual and trial-level factors did not have sufficient power to detect reliable differences in BMI z-score. Thus, the overall effectiveness of early obesity prevention interventions remains uncertain, as does whether there may be differential effects amongst subgroups.

### Need for individual participant data meta-analysis

The limitations and evidence gaps described above highlight the need for more powerful and in-depth analyses focusing on preventive interventions in very early childhood. Since the EPOCH PMA,^38^ we have identified more than 60 additional ongoing or completed very early obesity prevention trials worldwide with a combined sample size of more than 50,000 participants. While most trials are powered to detect some important differences in key outcomes, individually they have limited power to detect a difference in our primary outcome, BMI z-score at 24 +/- 6 months of age. In order to reliably detect a reduction in BMI z-score similar to that seen in EPOCH (−0.12),^38^ 2920 participants are required (90% power, two-sided 5% level of significance). Moreover, usually about four times that sample size (n∼12,000) is required to detect differences in subgroups.^48^ The expected total sample size for TOPCHILD will exceed these estimates (as by December 2020, 37 trials including 34,370 participants have already agreed to share their IPD).

Conducting a trial of this size would be time and resource intensive. A more efficient method is to combine IPD from trials in a pooled analysis to increase the sample size and therefore statistical power. This strengthens the chance of detecting intervention effect differences, and enables us to determine the size of such effects with greater certainty,^49^ while also allowing variation in study designs and population which heightens generalisability and allows a greater diversity to study effect modification for different subgroups of individuals or trial characteristics.^50^ Moreover, this collaborative approach maximises the use of existing data, thereby reducing research waste.

Thus, we will conduct an IPD meta-analysis with detailed subgroup analyses of all available trials to confirm whether early obesity prevention interventions commencing antenatally or in the first year after birth are effective, and whether effectiveness varies across subgroups defined by individual- or trial-level characteristics. The knowledge generated from this study can be used to inform decision-making around the design and implementation of more effective, efficient, equitable and targeted interventions for the prevention of childhood obesity and its sequelae.

### Objectives

This IPD meta-analysis will address the following research questions:

1. Compared with usual care/attentional control, what are the effects of parent/caregiver-focused behavioural obesity prevention interventions commencing during pregnancy or infancy on:

a. child BMI z-score at age 24 months (+/- 6 months)? (primary outcome)
b. child BMI z-score at alternative timepoints, other child weight-related measures, infant feeding, dietary intake, physical activity, sedentary behaviours, sleep, parenting measures and adverse events? (secondary outcomes)
2. Do intervention effects vary across individual-level characteristics (e.g. parental BMI, parity, socioeconomic position, birthweight)?
3. Do intervention effects vary across trial-level characteristics (e.g. access to existing well-child health care programs, intervention mode of delivery, timing of intervention onset)?

## METHODS AND ANALYSIS

We will conduct a systematic review with IPD meta-analysis and a nested PMA according to the methods recommended by the Cochrane Collaboration.^51, 52^ A nested PMA enables integration of prospective evidence into a retrospective meta-analysis, and harmonisation among planned/ongoing studies.^52^ Lead investigators of eligible trials will be invited to share their IPD and join the Transforming Obesity Prevention for CHILDren (TOPCHILD) Collaboration (www.topchildcollaboration.org). This protocol adheres to the Preferred Reporting Items for Systematic Reviews and Meta-Analysis extension for protocols (PRISMA-P)^53^ (Supplementary Appendix 1).

### Eligibility criteria

#### Types of studies

This systematic review will include randomised controlled trials only, including feasibility studies, pilot trials and definitive trials. Randomisation may occur at the individual level or by cluster (e.g. child care, community), including stepped-wedge designs. Quasi-randomised trials are excluded as they may introduce bias. There are no language or date restrictions.

#### Trial participants

Participants will be parents/caregivers (including pregnant women) and their infant(s) aged 0-12 months (at baseline). Caregiver is defined as the person with primary responsibility for care of the child, and excludes secondary sources of support, such as child care providers and early childhood teachers. Women may be primipara or multipara, and both singletons and multiples are eligible.

#### Types of interventions

Interventions must be behavioural interventions targeting parents/caregivers, and include at least one component related to modifiable child behaviours that may influence overweight/obesity risk (e.g. infant feeding, dietary intake, physical activity, sedentary behaviours, sleep). They may commence in the pre-conception or antenatal phase but must include intervention exposure targeting the birth to 12 months infancy stage, as pregnancy-only interventions are considered distinct and are currently being examined by Dodd et al in a separate IPD meta-analysis.^54^ Only childhood obesity prevention-focused trials will be included; these are defined as trials that clearly state childhood obesity prevention as a key aim/objective. Interventions focused only on improving an obesity related behaviour (e.g. sleep, delayed introduction of solid foods), as well as those focused on treatment of obesity, stunting or underweight will be excluded. Trials with a dual focus to prevent obesity and undernutrition are eligible, though we will carefully consider and pre-specify how their data will be incorporated in the statistical analysis plan. Interventions focused solely on nutritional supplements will be excluded, as they are not considered to be behavioural interventions.

#### Types of comparator/control

Eligible trials must have either 1) a usual care control arm, defined as existing local child health care, or 2) no intervention (including waitlist control) or 3) attention control (e.g. child safety education).

#### Types of outcome measures

To be included, trials must collect at least one of the child weight-related outcomes listed in Table 1 post intervention (at any age), i.e. BMI/BMI z-score, prevalence of overweight/obesity, percent fat content/adiposity, skinfold thickness, abdominal circumference, waist-to-height ratio. This is considered a legitimate and pragmatic approach given our review is of multi-component public health interventions focusing on obesity.^55^

**Table 1.** Outcomes and covariates/subgroups.

| ***Variable*** | ***Definition/explanatory text/examples*** |
| --- | --- |
| **Primary outcome** |  |
| BMI z-score at age 24 months (+/- 6 months) | determined in accordance with WHO growth standards<sup>60</sup> |
| **Secondary outcomes** |  |
| BMI z-score at 12 months (+/- 3 months) | determined in accordance with WHO growth standards<sup>60</sup> |
| BMI z-score at 48 months (+/- 12 months) | determined in accordance with WHO growth standards<sup>60</sup> |
| Other weight-related measures | e.g. prevalence of overweight/obesity (defined as BMI z-score of at least 2 standard deviations above the WHO reference), percent fat content/ adiposity, skinfold thickness, abdominal circumference, waist-to-height ratio, velocity of weight gain |
| Infant feeding | e.g. breastfeeding initiation and duration, exclusivity of breastfeeding, age at introduction of solid foods (complementary feeding) |
| Dietary intake | e.g. energy intake, intake of fruit, vegetables, energy dense nutrient poor (EDNP) foods, and sugar-sweetened beverages (SSBs) |
| Sedentary behaviours | e.g. screen time, restrained time while awake (in prams/strollers, high-chairs, strapped on a caregiver's back or chest) |
| Physical activity | e.g. active play duration, prone play ('tummy time'), device assessed physical activity time |
| Sleep | e.g. sleep duration, measures of sleep quality such as frequency and duration of |
|  | waking at night |
| Parental/caregiver measures | general and domain-specific parenting styles and practices, e.g. parenting self-efficacy, parenting styles, parent feeding practices, parent physical activity practices, parent sleep practices, stress |
| Adverse events | e.g. underweight, injuries, infection |
| **Individual-level covariates /subgroups** |  |
| Socioeconomic position | e.g. household income/country median household income, parent/caregiver highest education level, employment status |
| Parental weight status | e.g. maternal pre-pregnancy BMI, paternal BMI |
| Race/ethnicity | trialist defined |
| Maternal age | at recruitment |
| Maternal gestational weight gain | in kilograms |
| Parity | primipara, multiparous |
| Mode of delivery at birth | caesarean, vaginal |
| Birthweight | in grams |
| Weight for gestational age | small-for-gestational-age (SGA), appropriate-for-gestational-age (AGA), large-for-gestational-age (LGA) |
| Sex | female, male, uncertain/other |
| Gestational age at birth | preterm, term |
| Household composition | e.g. 2 vs 1 adult household, siblings, marital status |
| Type of pregnancy | singleton, multiple |
| Maternal diabetes | gestational, type 1, type 2 |
| Smoking during pregnancy | yes/no |
| Infant's age at enrolment | in months |
| Child's age at final assessment | in months |
| Child care attendance | yes/no |
| **Trial-level covariates/subgroups** |  |
| Delivery mode (intervention) | e.g. face-to-face, letter, mobile digital device, individual vs group |
| Intervention setting | e.g. household residence, community healthcare facility |
| Intervention dose/intensity | e.g. total number of contacts, frequency of contact, duration of contact |
| Fidelity | planned, actual |
| Timing of intervention onset | pre-conception, antenatal, postnatal |
| Timing of intervention completion | child age in months |
| Current level of background care in the community | descriptive, categorisation to be determined, e.g. expected number of health contacts between birth and 1 year, expectation of attending prenatal programs (yes/no), etc. |
| Country | low, middle, high income |
| Behavioural +/- other intervention type | behavioural intervention(s) alone vs behavioural + other intervention type (e.g. supplement) |

### Eligibility for nested prospective meta-analysis

In accordance with PMA methodology,^52^ only planned/ongoing trials will be eligible for the nested PMA if trial results were not yet known to the investigator/s at the time the main components of the TOPCHILD protocol (i.e. aims and objectives, hypotheses, eligibility criteria, main outcomes, subgroup and sensitivity analyses) were initially agreed in December 2020. We encourage investigators of planned/ongoing studies to collect the outcomes and covariates listed in Table 1 where possible, to facilitate data harmonisation and synthesis.

### Information sources and search strategy

In March 2020 we undertook an initial systematic search for eligible trials using the following databases from their inception: Medline (Ovid), Embase (Ovid), Cochrane Central Register of Controlled Trials (CENTRAL), CINAHL (EBSCO), PsycInfo, ClinicalTrials.gov and the World Health Organization’s International Clinical Trials Registry Platform’s (WHO ICTRP) Search Portal. The full search strategy is available in Supplementary Appendix 2. This search will be updated annually for the duration of the TOPCHILD Collaboration (currently funded until end 2023). Collaborators and contacts will also be asked to notify us of any planned, ongoing or completed trials of which they are aware that may meet the eligibility criteria.

#### Selection of studies for inclusion in the review

Two members of the TOPCHILD Steering Group will independently screen all retrieved records against eligibility criteria. Any discrepancies will be resolved by discussion or, if required, adjudication by a third reviewer from the Steering Group. The Principal Investigator and/or corresponding author of eligible trials will be invited by email to join the TOPCHILD Collaboration. If there is no response to initial emails and reminders, we will contact co-authors and/or other contacts listed in registration records and consult our existing networks to see if they can reach out to those they may know. If IPD cannot be obtained for an eligible trial, we will use aggregate data sourced from publications where available.

Supplementary Appendix 3 lists eligible trials identified up to March 2020.

### Data collection, management, and confidentiality

#### Data receipt / extraction

Trialists of all eligible studies will be invited to share de-identified individual participant data via secure data transfer platforms or via an institutional-secure email using password-protected zip files. Data will be provided according to a pre-specified coding template where possible. Otherwise, data will be accepted in any format and recoded as necessary. The data management team (within the TOPCHILD Steering Group) will receive and store the data in perpetuity in a secure, customised database at the NHMRC Clinical Trials Centre, University of Sydney, and data management will follow the *University of Sydney Data Management Policy 2014*. Each trial will also be asked to provide metadata (i.e. data that provides information about their trial dataset), such as questionnaires, data collection forms, and data dictionaries to aid understanding of the dataset. Trial-level data, such as setting, intervention timing, mode of delivery, comparator/control details, method of sequence generation, allocation concealment, geographical location, sample size, outcome measures and definitions will be extracted into a database and cross-checked against any published reports, trial protocols, registration records and data collection sheets.

#### Data processing

Data from each trial will be checked with respect to range, internal consistency, consistency with published reports and missing items. Integrity of the randomisation process will be examined by reviewing the chronological randomisation sequence and pattern of assignment, as well as the balance of participant characteristics across intervention and control groups. Any inconsistencies or missing data will be discussed with trialists and/or data managers and resolved by consensus. Once finalised, data from each of the trials will be combined into a single TOPCHILD Collaboration database.

### Risk of bias assessment and certainty of evidence appraisal

Included studies will be assessed for risk of bias by two independent reviewers from the TOPCHILD Steering Group using Version 2 of the Cochrane risk-of-bias tool for randomised trials (RoB 2).^56^ This tool includes five domains encompassing bias arising from: the randomisation process, deviations from intended interventions, missing outcome data, measurement of the outcome, and selection of the reported result. For cluster-randomised trials, bias arising from identification or recruitment of individual participants within clusters will also be assessed.^45^ The certainty of evidence will be assessed according to Cochrane procedures^57^ using the Grading of Recommendations Assessment, Development and Evaluation (GRADE) approach.^58^ Any differences will be resolved by consensus or with a third reviewer from the TOPCHILD Steering Group.

### Primary outcome

The primary outcome will be BMI z-score at age 24 months (+/- 6 months). BMI z-score will be determined in accordance with World Health Organization (WHO) growth standards.^47^

### Secondary outcomes

All outcomes are detailed in Table 1. Where possible, definitions will be standardised, otherwise outcomes will be used as defined within each trial. Secondary outcomes include BMI z-score at other timepoints, other measures of child weight, infant feeding (including breastfeeding and introduction of solid foods), dietary intake, sedentary behaviours, physical activity, and sleep, as well as parent/caregiver-related measures. We will also assess any adverse events, such as underweight or poor weight gain.

### Covariates/subgroups

All included covariates are listed in Table 1. Individual-level and trial-level subgroup analyses will be conducted for the primary outcome of BMI z-score at age 24 months (+/- 6 months). Those of primary interest at the individual level include socioeconomic position, race/ethnicity, pre-pregnancy maternal/paternal BMI, maternal age, gestational weight gain and parity, and at the trial level include timing of intervention onset, current level of background care in the community, recruitment country and mode of delivery.

Where possible, outcomes and covariates will be collected as continuous variables to maximise power to detect intervention effects and interactions, and enable exploration of any non-linear relationships.^59^ Dichotomous and categorical variables will also be collected to aid interpretation, and if data are insufficient for the pre-specified subgroup analyses, categories will be collapsed prior to any analyses being conducted.

### Data analysis

A detailed statistical analysis plan will be prepared and agreed upon by the TOPCHILD Collaboration members prior to any analyses being undertaken. Analyses will follow the intention-to-treat principle and include all randomised infant-parent/caregiver dyads for which data are available (including any that were excluded from the original study analysis). For cluster RCTs, correlated data will be taken into account by fitting the models using generalised estimating equations to derive appropriate standard errors. Correlations between multiples also will be accounted for in the analyses.

The primary analysis for all outcomes will be conducted using a one-stage approach combining all available IPD and aggregate data (where IPD are unavailable) to reduce the risk of availability bias.^61, 62^ The combined dataset will be analysed including trial as a random effect. Models will be chosen appropriate to the outcome type. Generalised linear models with appropriate distributions and link functions will be used for continuous and binary outcomes, while Cox proportional hazards regression will be used to analyse time-to-event outcomes subject to censoring. For example, linear models will be used for the primary outcome while relative risk binomial regression with log link function will be used for prevalence of overweight/obesity, and Cox models will be used for breastfeeding duration. Where possible, continuous covariates and outcomes will be analysed on their continuous scale to maximise utility of available data.^59^

Heterogeneity of intervention effects across trials will be investigated using quantitative measures (I^2^) supplemented by graphical presentations as recommended in the Cochrane Handbook.^63^ Any notable heterogeneity identified will be explored further to ascertain if the combination of trials is appropriate.

Results will be reported using appropriate estimates of intervention effect (relative risks, mean differences or hazard ratios) with 95% confidence intervals and associated two-sided p values. For trials with multiple intervention arms, we will present the data for each intervention arm compared to the control arm, with the number of participants in the control arm adjusted to ensure no double counting.^40^ Missing data will be explored in sensitivity analyses using appropriate methods. All analyses will be performed using the open-source software R.^64^

Differences in intervention effect between the pre-specified subgroups will be examined by testing a treatment by subgroup interaction term within the 1-stage-model. Findings of subgroup analyses will be reported as exploratory,^65^ and summarised using a 1-stage-approach supplemented by graphical presentation in a forest plot using a 2-stage-approach. Non-linear relationships will be explored for continuous covariates using a multivariate meta-analysis of the trend.^59^

Other exploratory analyses for the primary outcome will include graphical presentation of BMI z-score distributions to investigate any differences beyond mean differences and examine any non-linear relationships. The potential for mediation and moderator analyses using parent/caregiver measures will be explored and detailed in a statistical analysis plan after we have extracted information about relevant variables collected by included trials.

### Assessment of selection or publication bias

Potential selection bias and publication bias will be investigated by conducting a nested prospective meta-analysis (PMA) and comparing prospectively versus retrospectively included trials in a sensitivity analysis.^52^ We will also seek to include any unreported outcomes sourced from each trial’s IPD, which may alleviate selective outcome reporting bias.^51^ Lastly, contour-enhanced funnel plots will be used to examine whether there are differences in results between more and less precise studies.

### Adjustments for multiple testing

Only one primary outcome was selected for this study (Table 1). For secondary outcomes and subgroup analyses, no formal adjustments will be made for the potential inflation of type 1 error rates due to multiple testing. Instead, we will follow Schulz and Grimes’ approach^54^ and recommendations of the Cochrane Collaboration.^63^ This involves cautious interpretation of the magnitudes of effect, patterns and consistency of results across related outcomes and clinical/biological plausibility rather than focusing on any single statistically significant result in isolation which can be extremely misleading.^63, 65^

### Planned sensitivity analyses

Where possible, the following sensitivity analyses will be conducted for the primary outcome:

- Two stage approach
- Including IPD only, i.e. excluding trials without IPD available^55^
- Including prospectively included trials only (nested PMA), i.e. planned/ongoing trials for which results were not yet known to investigator/s at the time the main components of the TOPCHILD protocol were agreed^52^
- Adjusting for birthweight as a covariate
- Excluding trials with a high risk of bias for sequence generation and/or allocation concealment and/or loss to follow-up
- Excluding trials with a significant conflict of interest (e.g. funded by industry)
- The impact of missing data on conclusions about the intervention effect (if appropriate)

### Project management

Membership of the TOPCHILD Collaboration includes trial representatives from each of the trials contributing IPD to the project, a Steering Group and an Advisory Group. Trial representatives have the opportunity to contribute their expert knowledge to the TOPCHILD Collaboration and provide input into the protocols, statistical analysis plan, and final results manuscript. The Steering Group will be responsible for data collection, management and analysis, as well as communication within the Collaboration, including newsletter updates, maintenance of the TOPCHILD website and organisation of virtual or face-to-face collaborator meetings. The Advisory Group will comprise invited experts in childhood obesity prevention, IPD meta-analysis, statistics, behaviour change theory/methods and policy implementation.

### Patient and public involvement

The TOPCHILD Collaboration involves a broad range of stakeholders including health professionals, policy-makers and trialists. In addition, the Advisory Group includes a parent representative and intervention facilitator/nurse. They have given input into and feedback on this protocol and will be involved in discussion and interpretation of results.

## ETHICS AND DISSEMINATION

### Ethical considerations

IPD will be provided by each included trial on the stipulation that ethical approval has been provided by their respective Human Research Ethics Committees (or equivalent), and participants gave informed consent before enrolment to participate in the initial individual trials. Trialists remain the custodians of their own data, which will be de-identified before being shared with the TOPCHILD Collaboration. Ethical approval for this project has been granted by The University of Sydney Human Research Ethics Committee (2020/273) and Flinders University Social and Behavioural Research Ethics Committee (project no. HREC CIA2133-1).

### Publication policy

TOPCHILD manuscripts will be prepared by the Steering Group in consultation with the Advisory Group, and circulated to the full Collaboration for comment, revision and approval prior to submission for publication. Any reports of the results of this study will be published either in the name of the collaborative group, or by representatives of the collaborative group on behalf of the TOPCHILD Collaboration, as agreed by members of the collaborative group.

## DISCUSSION

This will be the largest IPD meta-analysis to date of trials evaluating behavioural obesity prevention interventions commencing in very early childhood. The findings will inform next generation obesity prevention initiatives that are effective, efficient, and equitable. Such interventions could set children on a better health trajectory early on and reduce the potentially life-long burden of disease associated with obesity.

The main strengths of this study arise from use of IPD meta-analysis methodology, which is considered the ‘gold standard’.^66^ It involves collecting the raw line-by-line data for each participant in each study from the original trialists. This can improve the quality of data, and enables more in-depth and precise analyses than would be possible using only published aggregate data.^51^ In particular, IPD meta-analysis will enable thorough exploration of individual-level and trial-level subgroups, so that we may quantify any differential effects and uncover the key determinants of successful outcomes. This addresses the limitations identified in previous reviews of childhood obesity prevention,^38, 40, 45^ where such detailed and sufficiently powered analyses were simply not possible.

A potential limitation of this study is the risk of not obtaining IPD from all eligible studies, resulting in inclusion bias. Where available, we will include aggregate data from these studies, and conduct sensitivity analyses with inclusion of IPD only to explore potential bias.^67^ Further, there may be variations across studies in how measures are collected and reported, which may lead to some imprecision and difficulties pooling the data. We will seek to address this using nested prospective meta-analysis methodology, whereby researchers of planned or ongoing trials are encouraged to harmonise their trial design and collect core outcome measures to facilitate meta-analysis and interpretation.^52^ For completed studies, we will derive common outcome variables by cleaning, re-coding, and converting existing measures where possible.

We plan to complete the first round of study identification and IPD collection by early 2021, then conduct the primary analyses and disseminate the results by the end of 2022. Trials that are not completed in time to provide data for this cycle will remain a part of the TOPCHILD Collaboration, and their data will be included in future updates of TOPCHILD.

This IPD meta-analysis will be conducted in parallel with a complementary TOPCHILD project (Johnson et al, unpublished), which aims to deconstruct childhood obesity interventions into their components (i.e. delivery features, target behaviours and behaviour change techniques) using systematic, internationally recognised frameworks and both published and unpublished trial materials. In future, the resulting dataset curated from these two projects will be used for predictive modelling of intervention component effectiveness at an individual participant level, facilitating a personalised or precision medicine approach to public health prevention.

The TOPCHILD Collaboration will maximise use of existing trial data that will enable us to understand and utilise the most effective intervention components for specific population groups and contexts. It will provide urgently needed evidence to inform development and implementation of effective, efficient and equitable interventions for the prevention of early childhood obesity. The results will be of prime importance for guideline developers, policymakers, consumers and the research community. Further information and updates on the TOPCHILD Collaboration can be found at www.topchildcollaboration.org.

## Data Availability

No data are available for the current manuscript since this is a protocol.

## Acknowledgements

The authors thank Slavica Berber for guidance and input into the database search strategy. We would also like to acknowledge the NHMRC Centre of Research Excellence in Early Prevention of Obesity in Childhood (EPOCH CRE), who supported the pilot and foundational work for this project. Lastly, we thank Maria Luisa Garmendia and Camila Corvalan for their feedback on an earlier version of this protocol.

TOPCHILD Collaboration members

<u>Steering group:</u> Anna Lene Seidler, Kylie Hunter, Brittany Johnson, Rebecca Golley, Angie Barba, Lisa Askie, Mason Aberoumand, Sol Libesman

<u>Advisory group:</u> Louise Baur, Paul Chadwick, David Espinoza, Alison Hayes, Ian Marschner, Seema Mihrshahi, Denise O’Connor, Chris Rissel, Kristy Robledo, Lee Sanders, Wendy Smith, Lukas Staub, Michelle Sue-See, Sarah Taki, Rachael Taylor, Luke Wolfenden, Charles Wood, Shonna Yin

<u>Trial representatives (to date):</u> Stephanie Anzman-Frasca, Jinan Banna, Maria Bryant, Karen Campbell, Maribel Campos Rivera, Eva Corpeleijn, Lynne Daniels, Kayla de la Haye, Ata Ghaderi, Carolina González Acero, Michael Goran, Rachel Gross, Christine Helle, Kylie Hesketh, Kaumudi Joshipura, Alison Karasz, Levie Karssen, Rajalakshmi Lakshman, Junilla Larsen, Ana Maria Linares, Claudio Maffeis, Mary Jo Messito, Emily Oken, Ken Ong, Sharleen O’Reilly, Nina Øverby, Cristina Palacios, Ian Paul, Ana Perez-Exposito, Eliana Perrin, Hein Raat, Finn Rasmussen, Margrethe Røed, Russell Rothman, Sarah-Jeanne Salvy, Jennifer Savage, Cathleen Odar Stough, Barry Taylor, Rachael Taylor, Amanda Thompson, Jessica Thomson, Vera Verbestel, Heather Wasser, Li Ming Wen

## Author contributions

ALS together with KEH, BJJ, LA, RKG conceived the idea for the study.

KEH, BJJ, ALS, LA, RKG developed the research question and protocol registration. KEH wrote the first draft of the manuscript.

KEH, BJJ, ALS, RKG, LA developed the eligibility criteria and search strategy. KEH, MA, AB, BJJ, SL, ALS performed the search and screening.

AJH, CR, CTW, DE, DAO’C, ICM, KPR, LAB, LPS, LW, MS-S, PC, RWT, SM, ST, WS provided critical review and feedback at each stage of the process.

All authors critically revised the manuscript for intellectual content, and agreed and approved the final manuscript. KEH is the guarantor of the review.

