## Supplementary File 2 for "Transforming Obesity Prevention for CHILDren (TOPCHILD) Collaboration: protocol for a systematic review with individual participant data meta-analysis of behavioural interventions for the prevention of early childhood obesity"

Supplementary file 2: TOPCHILD search strategy

### Ovid MEDLINE(R)

1. pediatric obesity/
2. Weight Gain/
3. obes*.ti,ab
4. (weight gain).ti,ab
5. (overweight or over weight).ti,ab
6. weight change*.ti,ab
7. ((bmi or body mass index) adj2 (gain or loss or change)).ti,ab
8. 1 or 2 or 3 or 4 or 5 or 6 or 7
9. social support/
10. ((behaviour or behavior) and change).ti,ab
11. ((behavio?r*) adj (therapy or modif* or strateg* or intervention* or advice or program* or class* or counsel* or educat* or instruct* or teach* or train* or guidance or lesson* or workshop* or module* or consultation* or session*)).ti,ab
12. ((lifestyle or life style) adj (chang* or modif* or strateg* or intervention* or advice or program* or class* or counsel* or educat* or instruct* or teach* or train* or guidance or lesson* or workshop* or module* or consultation* or session*)).ti,ab
13. social support.ti,ab
14. (peer adj2 support).ti,ab
15. counsel?ing.ti,ab
16. education* adj1 (intervention* or program* or class* or counsel* or teach* or workshop* or module* or consultation* or session*)).ti,ab
17. home visit*.ti,ab
18. 9 or 10 or 11 or 12 or 13 or 14 or 15 or 16 or 17
19. exp Breastfeeding/
20. Infant Nutritional Physiological Phenomena/
21. Child Nutrition Sciences/
22. Infant Food/
23. ((child or toddler or infant$) adj1 (food or feeding or nutrition$)).tw.
24. ((responsive or complementary) adj1 feeding).ti,ab
25. ((diet* or nutrition) adj (modif* or strateg* or intervention* or advice or program* or class* or counsel* or educat* or instruct* or teach* or train* or guidance or lesson* or workshop* or module* or consultation* or session*)).ti,ab
26. (healthy eating).ti,ab
27. (fruit or vegetable*).ti,ab
28. (high fat* or low fat* or fatty food*).ti,ab
29. 19 or 20 or 21 or 22 or 23 or 24 or 25 or 26 or 27 or 28
30. exp Exercise/
31. exercis*.ti,ab
32. (physical activity or physical inactivity).ti,ab
33. sedentary behavio?r.ti,ab
34. (screen time).ti,ab
35. 30 or 31 or 32 or 33 or 34
36. Sleep/
37. Exp Primary prevention
38. exp Health Promotion/
39. exp Health Education/
40. prevention.mp
41. prevent*.ti,ab
42. (health promotion or health education or health communication).ti,ab
43. exp Obesity/pc (Prevention and Control)
44. exp Overweight/pc (Prevention & Control)
45. (obesity adj2 prevent*).ti,ab
46. (overweight adj2 prevent*).ti,ab
47. 37 or 38 or 39 or 40 or 41 or 42 or 43 or 44 or 45 or 46
48. 8 and (18 or 29 or 35 or 36) and 47
49. exp child/ or exp infant/
50. ((child* or infant* or baby or toddler* or pediatr* or paediatr*) not adolescen*).ti,ab
51. (pregnan* or antenatal or parent or parent$1 or care giver or caregiver or guardian or family or families or mother$1 or father$1).ti,ab
52. 49 or 50 or 51
53. 48 and 52
54. (exp animals/ not humans.sh.) or (rat or rats or mouse or mice or rodent*).ti.
55. 53 not 54
56. controlled clinical trial.pt.
57. randomi#ed.ti,ab.
58. randomly.ab.
59. (clinical trials as topic or controlled clinical trials as topic).sh.
60. trial.ti.
61. exp randomized controlled trial/ or exp randomized controlled trials as topic/
62. 56 or 57 or 58 or 59 or 60 or 61
63. 55 and 62

### Embase Classic+Embase

1. pediatric obesity/
2. Weight Gain/
3. obes*.ti,ab
4. (weight gain).ti,ab
5. (overweight or over weight).ti,ab
6. weight change*.ti,ab
7. ((bmi or body mass index) adj2 (gain or loss or change)).ti,ab
8. 1 or 2 or 3 or 4 or 5 or 6 or 7
9. social support/
10. ((behviour or behavior) and change).ti,ab
11. ((behavio?r*) adj (therapy or modif* or strateg* or intervention* or advice or program* or class* or counsel* or educat* or instruct* or teach* or train* or guidance or lesson* or workshop* or module* or consultation* or session*)).ti,ab
12. ((lifestyle or life style) adj (chang* or modif* or strateg* or intervention* or advice or program* or class* or counsel* or educat* or instruct* or teach* or train* or guidance or lesson* or workshop* or module* or consultation* or session*)).ti,ab
13. social support.ti,ab
14. (peer adj2 support).ti,ab
15. counsel?ing.ti,ab
16. education* adj1 (intervention* or program* or class* or counsel* or teach* or workshop* or module* or consultation* or session*)).ti,ab
17. home visit*.ti,ab
18. 9 or 10 or 11 or 12 or 13 or 14 or 15 or 16 or 17
19. exp Breastfeeding/
20. Infant Nutritional Physiological Phenomena/
21. Child Nutrition Sciences/
22. Infant Food/
23. ((child or toddler or infant$) adj1 (food or feeding or nutrition$)).tw.
24. ((responsive or complementary) adj1 feeding).ti,ab
25. ((diet* or nutrition) adj (modif* or strateg* or intervention* or advice or program* or class* or counsel* or educat* or instruct* or teach* or train* or guidance or lesson* or workshop* or module* or consultation* or session*)).ti,ab
26. (healthy eating).ti,ab
27. (fruit or vegetable*).ti,ab
28. (high fat* or low fat* or fatty food*).ti,ab
29. 19 or 20 or 21 or 22 or 23 or 24 or 25 or 26 or 27 or 28
30. exp Exercise/
31. exercis*.ti,ab
32. (physical activity or physical inactivity).ti,ab
33. sedentary behavio?r.ti,ab
34. (screen time).ti,ab
35. 30 or 31 or 32 or 33 or 34
36. Sleep/
37. Exp Primary prevention
38. exp Health Promotion/
39. exp Health Education/
40. prevention.mp
41. prevent*.ti,ab
42. (health promotion or health education or health communication).ti,ab
43. exp Obesity/pc (Prevention and Control)
44. exp Overweight/pc (Prevention & Control)
45. (obesity adj2 prevent*).ti,ab
46. (overweight adj2 prevent*).ti,ab
47. 37 or 38 or 39 or 40 or 41 or 42 or 43 or 44 or 45 or 46
48. 8 and (18 or 29 or 35 or 36) and 47
49. exp child/ or exp infant/
50. (child* or infant* or baby or toddler* or pediatr* or paediatr* not adolescen*).ti,ab
51. (pregnan* or antenatal or parent or parent$1 or care giver or caregiver or guardian or family or families or mother$1 or father$1).ti,ab
52. 49 or 50 or 51
53. 48 and 52
54. (exp animals/ not humans.sh.) or (rat or rats or mouse or mice or rodent*).ti.
55. 53 not 54
56. exp clinical trial/
57. exp Randomized Controlled Trial/
58. randomization/
59. clinical trial.tw.
60. random$.tw.
61. Comparative Study/
62. clinical trial.tw.
63. (comparison group$ or control group$).tw.
64. 56 or 57 or 58 or 59 or 60 or 61 or 62 or 63
65. 55 and 64

### EBM Reviews - Cochrane Central Register of Controlled Trials

1. pediatric obesity/
2. Weight Gain/
3. obes*.ti,ab
4. (weight gain).ti,ab
5. (overweight or over weight).ti,ab
6. weight change*.ti,ab
7. ((bmi or body mass index) adj2 (gain or loss or change)).ti,ab
8. 1 or 2 or 3 or 4 or 5 or 6 or 7
9. social support/
10. ((behviour or behavior) and change).ti,ab
11. ((behavio?r*) adj (therapy or modif* or strateg* or intervention* or advice or program* or class* or counsel* or educat* or instruct* or teach* or train* or guidance or lesson* or workshop* or module* or consultation* or session*)).ti,ab
12. ((lifestyle or life style) adj (chang* or modif* or strateg* or intervention* or advice or program* or class* or counsel* or educat* or instruct* or teach* or train* or guidance or lesson* or workshop* or module* or consultation* or session*)).ti,ab
13. social support.ti,ab
14. (peer adj2 support).ti,ab
15. counsel?ing.ti,ab
16. education* adj1 (intervention* or program* or class* or counsel* or teach* or workshop* or module* or consultation* or session*)).ti,ab
17. home visit*.ti,ab
18. 9 or 10 or 11 or 12 or 13 or 14 or 15 or 16 or 17
19. exp Breast feeding/
20. Infant Nutritional Physiological Phenomena/
21. Child Nutrition Sciences/
22. Infant Food/
23. ((child or toddler or infant$) adj1 (food or feeding or nutrition$)).tw.
24. ((responsive or complementary) adj1 feeding).ti,ab
25. ((diet* or nutrition) adj (modif* or strateg* or intervention* or advice or program* or class* or counsel* or educat* or instruct* or teach* or train* or guidance or lesson* or workshop* or module* or consultation* or session*)).ti,ab
26. (healthy eating).ti,ab
27. (fruit or vegetable*).ti,ab
28. (high fat* or low fat* or fatty food*).ti,ab
29. 19 or 20 or 21 or 22 or 23 or 24 or 25 or 26 or 27 or 28
30. exp Exercise/
31. exercis*.ti,ab
32. (physical activity or physical inactivity).ti,ab
33. sedentary behavio?r.ti,ab
34. (screen time).ti,ab
35. 30 or 31 or 32 or 33 or 34
36. Sleep/
37. Exp Primary prevention
38. exp Health Promotion/
39. exp Health Education/
40. prevention.mp
41. prevent*.ti,ab
42. (health promotion or health education or health communication).ti,ab
43. (obesity adj2 prevent*).ti,ab
44. (overweight adj2 prevent*).ti,ab
45. 37 or 38 or 39 or 40 or 41 or 42 or 43 or 44
46. 8 and (18 or 29 or 35 or 36) and 45
47. exp child/ or exp infant/
48. (child* or infant* or baby or toddler* or pediatr* or paediatr* not adolescen*).ti,ab
49. (pregnan* or antenatal or parent or parent$1 or care giver or caregiver or guardian or family or families or mother$1 or father$1).ti,ab
50. 49 or 50 or 51
51. 46 and 50
52. (exp animals/ not humans.sh.) or (rat or rats or mouse or mice or rodent*).ti.
53. 51 not 52
54. 51 and 53

### CINAHL

| S70 | S60 AND S69 |
| --- | --- |
| S69 | S61 OR S62 OR S63 OR S64 OR S65 OR S66 OR S67 OR S68 |
| S68 | TX comparison group* |
| S67 | TX random* |
| S66 | "clin* N25 trial*" |
| S65 | (PT "CLINICAL TRIAL") |
| S64 | (MH "Clinical Trials") |
| S63 | (MH "Random Sample+") |
| S62 | (MH "Random Assignment") |
| S61 | (MH "Comparative Studies") |
| S60 | S58 NOT S59 |
| S59 | (MH "Animals+") |
| S58 | S55 AND S57 |
| S57 | S10 AND S49 AND S56 |
| S56 | S21 OR S34 OR S40 OR S41 |
| S55 | S50 OR S51 OR S52 OR S53 OR S54 |
| S54 | (TI pregnan* or antenatal or parent* or care giver or caregiver or guardian or family or families or mother$1 or father$1) OR (AB pregnan* or antenatal or parent* or care giver or caregiver or guardian or family or families or mother$1 or father$1) |
| S53 | (TI child* or infant* or baby or toddler* or pediatr* or paediatr* not adolescen*) OR (AB child* or infant* or baby or toddler* or pediatr* or paediatr* not adolescen*) |
| S52 | (MH "Infant+") |
| S51 | (MH "Child") |
| S50 | (MH "Child+") |
| S49 | S42 OR S43 OR S44 OR S45 OR S46 OR S47 OR S48 |
| S48 | (TI overweight N2 prevent*) OR (AB overweight N2 prevent*) |
| S47 | (TI obesity N2 prevent*) OR (AB obesity N2 prevent*) |
| S46 | (TI health promotion or health education or health communication) OR (AB health promotion or health education or health communication) |
| S45 | (TI prevent*) OR (AB prevent*) |
| S44 | (MH "Health Education+") |
| S43 | (MH "Health Promotion+") |
| S42 | "primary prevention" |
| S41 | (MH "Sleep+") |
| S40 | S35 OR S36 OR S37 OR S38 OR S39 |
| S39 | (TI screen time) OR (AB screen time) |
| S38 | (TI sedentary behavio?r) or (AB sedentary behavio?r) |
| S37 | (TI physical activity or physical inactivity) OR (AB physical activity or physical inactivity) |
| S36 | (TI exercis*) OR (AB exercis*) |
| S35 | (MH "Exercise+") |
| S34 | S22 OR S23 OR S24 OR S25 OR S26 OR S27 OR S28 OR S29 OR S30 OR S31 OR S32 OR S33 |
| S33 | (TI high fat* or low fat* or fatty food*) OR (AB high fat* or low fat* or fatty food*) |
| S32 | (TI fruit or vegetable*) OR (AB fruit or vegetable*) |
| S31 | (TI healthy eating) OR (AB healthy eating) |
| S30 | (AB nutrition N2 modif*) or (AB nutrition N2 strateg*) or (AB nutrition N2 intervention*) or (AB nutrition N2 advice) or (AB nutrition N2 program*) or (AB nutrition N2 class*) or (AB nutrition N2 counsel*) or (AB nutrition N2 educat*) or (AB nutrition N2 instruct*) or (AB nutrition N2 teach*) or (AB nutrition N2 train*) or (AB nutrition N2 guidance) or (AB nutrition N2 lesson*) or (AB nutrition N2 workshop*) or (AB nutrition N2 module*) or (AB nutrition N2 consultation*) or (AB nutrition N2 session*) |
| S29 | (AB diet* N2 modif*) or (AB diet* N2 strateg*) or (AB diet* N2 intervention*) or (AB diet* N2 advice) or (AB diet* N2 program*) or (AB diet* N2 class*) or (AB diet* N2 counsel*) or (AB diet* N2 educat*) or (AB diet* N2 instruct*) or (AB diet* N2 teach*) or (AB diet* N2 train*) or (AB diet* N2 guidance) or (AB diet* N2 lesson*) or (AB diet* N2 workshop*) or (AB diet* N2 module*) or (AB diet* N2 consultation*) or (AB diet* N2 session*) |
| S28 | (TI nutrition N2 modif*) or (TI nutrition N2 strateg*) or (TI nutrition N2 intervention*) or (TI nutrition N2 advice) or (TI nutrition N2 program*) or (TI nutrition N2 class*) or (TI nutrition N2 counsel*) or (TI nutrition N2 educat*) or (TI nutrition N2 instruct*) or (TI nutrition N2 teach*) or (TI nutrition N2 train*) or (TI nutrition N2 guidance) or (TI nutrition N2 lesson*) or (TI nutrition N2 workshop*) or (TI nutrition N2 module*) or (TI nutrition N2 consultation*) or (TI nutrition N2 session*) |
| S27 | (TI diet* N2 modif*) or (TI diet* N2 strateg*) or (TI diet* N2 intervention*) or (TI diet* N2 advice) or (TI diet* N2 program*) or (TI diet* N2 class*) or (TI diet* N2 counsel*) or (TI diet* N2 educat*) or (TI diet* N2 instruct*) or (TI diet* N2 teach*) or (TI diet* N2 train*) or (TI diet* N2 guidance) or (TI diet* N2 lesson*) or (TI diet* N2 workshop*) or (TI diet* N2 module*) or (TI diet* N2 consultation*) or (TI diet* N2 session*) |
| S26 | (MH "Infant Feeding+") |
| S25 | (MH "Infant Food+") |
| S24 | (MH "Child Nutritional Physiology+") |
| S23 | (MH "Infant Nutritional Physiology+") |
| S22 | (MH "Breast Feeding+") |
| S21 | S11 OR S12 OR S13 OR S14 OR S15 OR S16 OR S17 OR S18 OR S19 OR S20 |
| S20 | (TI home visit) OR (AB home visit) |
| S19 | (TI education N2 intervention*) OR (TI education N2 program*) OR (TI education N2 class*) OR (TI education N2 counsel*) OR (TI education N2 teach*) OR (TI education N2 workshop) OR (TI education N2 module*) OR (TI education N2 consultation*) OR (TI education N2 session*) (AB education N2 intervention*) OR (AB education N2 program*) OR (AB education N2 class*) OR (AB education N2 counsel*) OR (AB education N2 teach*) OR (AB education N2 workshop) OR (AB education N2 module*) OR (AB education N2 consultation*) OR (AB education N2 session*) |
| S18 | (TI counselling or counseling) OR (AB counselling or counseling) |
| S17 | (TI peer N2 support) OR (AB peer N2 support) |
| S16 | (TI social support) OR (AB social support) |
| S15 | (MM "Life Style Changes") |
| S14 | (MM "Behavior Modification+") |
| S13 | (MM "Behavioral Changes") |
| S12 | (TI behaviour change) OR (AB behaviour change) OR (TI behavior change) OR (AB behavior change) |
| S11 | (MH "Support, Psychosocial") |
| S10 | S1 OR S2 OR S3 OR S4 OR S5 OR S6 OR S7 OR S8 OR S9 |
| S9 | (TI body mass index N2 loss) OR (AB body mass index N2 loss) |
| S8 | (TI body mass index N2 gain) OR (AB body mass index N2 gain) |
| S7 | (TI body mass index N2 change) OR (AB body mass index N2 change) |
| S6 | (TI weight change*) OR (AB weight change*) |
| S5 | (TI overweight or over weight) OR (AB overweight or over weight) |
| S4 | (TI weight gain) OR (AB weight gain) |
| S3 | (TI obese or obesity) OR (AB obese or obesity) |
| S2 | (MH "Weight Gain") |
| S1 | (MH "Pediatric Obesity+") |

### APA PsycInfo

| 1 | exp obesity/ |
| --- | --- |
| 2 | exp overweight/ |
| 3 | Weight Gain/ |
| 4 | obes*.ti,ab. |
| 5 | weight gain.ti,ab. |
| 6 | (overweight or over weight).ti,ab. |
| 7 | weight change*.ti,ab. |
| 8 | Body Mass Index/ |
| 9 | ((bmi or "body mass index") adj2 (gain or loss or change)).ti,ab. |
| 10 | 1 or 2 or 3 or 4 or 5 or 6 or 7 or 8 or 9 |
| 11 | social support/ |
| 12 | ((behaviour or behavior) and change).ti,ab. |
| 13 | (behavio?r* adj (therapy or modif* or strateg* or intervention* or advice or program* or class* or counsel* or educat* or instruct* or teach* or train* or guidance or lesson* or workshop* or module* or consultation* or session*)).ti,ab. |
| 14 | ((lifestyle or life style) adj (chang* or modif* or strateg* or intervention* or advice or program* or class* or counsel* or educat* or instruct* or teach* or train* or guidance or lesson* or workshop* or module* or consultation* or session*)).ti,ab. |
| 15 | social support.ti,ab. |
| 16 | (peer adj2 support).ti,ab. |
| 17 | counsel?ing.ti,ab. |
| 18 | (education* adj1 (intervention* or program* or class* or counsel* or teach* or workshop* or module* or consultation* or session*)).ti,ab. |
| 19 | home visit*.ti,ab. |
| 20 | 11 or 12 or 13 or 14 or 15 or 16 or 17 or 18 or 19 |
| 21 | exp Breast feeding/ |
| 22 | Nutrition/ |
| 23 | Eating Behavior/ |
| 24 | ((child or toddler or infant$) adj1 (food or feeding or nutrition$)).tw. |
| 25 | ((responsive or complementary) adj1 feeding).ti,ab. |
| 26 | ((diet* or nutrition) adj (modif* or strateg* or intervention* or advice or program* or class* or counsel* or educat* or instruct* or teach* or train* or guidance or lesson* or workshop* or module* or consultation* or session*)).ti,ab. |
| 27 | healthy eating.ti,ab. |
| 28 | (fruit or vegetable*).ti,ab. |
| 29 | (high fat* or low fat* or fatty food*).ti,ab. |
| 30 | 21 or 22 or 23 or 24 or 25 or 26 or 27 or 28 or 29 |
| 31 | exp Exercise/ |
| 32 | exercis*.ti,ab. |
| 33 | (physical activity or physical inactivity).ti,ab. |
| 34 | sedentary behavio?r.ti,ab. |
| 35 | screen time.ti,ab. |
| 36 | 31 or 32 or 33 or 34 or 35 |
| 37 | sleep/ |
| 38 | exp prevention/ |
| 39 | exp Health Promotion/ |
| 40 | exp Health Education/ |
| 41 | prevention.mp. |
| 42 | prevent*.ti,ab. |
| 43 | (health promotion or health education or health communication).ti,ab. |
| 44 | (obesity adj2 prevent*).ti,ab. |
| 45 | (overweight adj2 prevent*).ti,ab. |
| 46 | 38 or 39 or 40 or 41 or 42 or 43 or 44 or 45 |
| 47 | 10 and (20 or 30 or 36 or 37) and 46 |
| 48 | ((child* or infant* or baby or toddler* or pediatr* or paediatr*) not adolescen*).ti,ab. |
| 49 | (pregnan* or antenatal or parent or parent$1 or care giver or caregiver or guardian or family or families or mother$1 or father$1).ti,ab. |
| 50 | 48 or 49 |
| 51 | 47 and 50 |
| 52 | (exp animals/ not humans.sh.) or (rat or rats or mouse or mice or rodent*).ti. |
| 53 | 51 not 52 |
| 54 | exp experimental design/ |
| 55 | randomi#ed.ti,ab. |
| 56 | randomly.ab. |
| 57 | exp clinical trials/ |
| 58 | trial.ti. |
| 59 | exp randomized controlled trial/ or exp randomized controlled trials as topic/ |
| 60 | 54 or 55 or 56 or 57 or 58 or 59 |
| 61 | 53 and 60 |

### WHO ICTRP

**Search string**

Basic search

1. infant AND obesity prevention

2. infant AND prevention of obesity

3. infant AND overweight prevention

4. infant AND prevention of overweight

5. infant AND prevent AND obesity

6. child AND obesity prevention

7. child AND prevention of obesity

8. child AND overweight prevention

9. child AND prevention of overweight

10. child AND prevent AND obesity

Advanced search

1. Title: prevent AND obesity

Recruitment Status: All

Limit: Search for clinical trials in children

2. Title: prevent AND overweight

Recruitment Status: All

Limit: Search for clinical trials in children

3. Title: prevent OR prevention

Condition: obesity OR overweight

Recruitment Status: All

Limit: Search for clinical trials in children

### Clinicaltrials.gov

**Search string**

Basic search

1. Condition: Obesity, Childhood

Other Terms: prevention

2. Condition: Obesity, Childhood

Other Terms: prevent
