## Supplementary File 3 for "Transforming Obesity Prevention for CHILDren (TOPCHILD) Collaboration: protocol for a systematic review with individual participant data meta-analysis of behavioural interventions for the prevention of early childhood obesity"

**Supplementary file 3: Eligible randomised controlled trials for the Transforming Obesity Prevention for Children (TOPCHILD) Collaboration**

*This table shows all trials that we have identified up to March 2020 as being eligible for inclusion in TOPCHILD. Trials marked in blue have already agreed to join the Collaboration and share their data and unpublished intervention materials.*

| **Trial Country/ies (PI)** | **Registration number (registration year)** | **Protocol publication year** | **Main results publication year** | **Start year/ completion year** | **Sample size** | **Participants** | **Primary outcome/s** |
| --- | --- | --- | --- | --- | --- | --- | --- |
| Australia  (Campbell 2013)^1^ | ISRCTN81847050  (2008) | 2008^2^ | 2013^1^ | 2008/2010 | 542 | First-time parent regularly attending first-time parent group | Dietary intake |
| Australia  (Campbell 2016)^3^ | ACTRN12611000386932  (2011) | 2016^3^ | na | 2011/na | 560 | First-time parent groups with children aged 3-4 months | Anthropometry: height, weight, waist circumference, BMI z-score at 18 and 36 months of age |
| Australia  (Daniels)^4^ | ACTRN12608000056392  (2008) | 2009^5^ | 2013^4^ | 2008/2009 | 698 | First-time mothers of healthy term infants | Food intake, food preferences, and feeding behaviour |
| Australia  (Wen 2007)^6^ | ACTRN12607000168459  (2007) | 2007^7^ | 2012^6^ | 2007/2010 | 782 | Women expecting their first child, between 24-34 weeks pregnancy | BMI at 2 years of age |
| Australia  (Wen 2019)^8^ | ACTRN12616001470482  (2016) | 2019^9^ | 2020^8^ | 2017/2020 | 1150 | Women in their third trimester | BMI, breastfeeding duration, and timing of introduction of solids |
| Belarus  (Oken/Kramer 2009)^10^ | ISRCTN37687716  (2005)  NCT01561612  (2012) | na | 2009^10^ | 1996/1998 | 17,046 | Mother-infant pairs consisting of full-term singleton infants weighing at least 2500 g and their healthy mothers who intended to breastfeed | Duration of any breastfeeding, prevalence of predominant  & exclusive breastfeeding at 3 & 6 months of life, occurrence of 1 or more episodes of gastrointestinal tract infection, 2 or more episodes of respiratory tract infection, and atopic eczema during the first 12 months of life |
| Belgium  (Verbestel)^11^ | na | na | 2013^11^ | 2009/2009 | 203 | Parents of children aged 9–24 months | BMI z-score, children’s lifestyle behaviours |
| Brazil  (Vitolo)^12^ | NCT00629629  (2008) | na | 2010^12^ | 2001/2003 | 500 | Newborn infants with birthweight ≥ 2500 g and gestational age ≥ 37 weeks | Exclusive breastfeeding at 1 year, lipid profile at 7-8 years of age |
| Canada  (Dennis)^13^ | ISRCTN13308752  (2019) | na | na | 2019/2022 | 5230 | Non-pregnant mother with no child or one child between 3-12 months | Proportion of infants with a BMI>85 percentile using World Health Organization growth reference range by sex and measured at 5 years of age |
| China  (Huang)^14^ | ChiCTR1800017773  (2018) | na | na | 2018/2029 | 4500 | Women between 20-42 years of age, planning pregnancy in the next 6 months or <14 weeks pregnant, singleton pregnancy | Obesity and overweight at 5 years of age |
| Denmark  (Pryds)^15^ | NCT01235663  (2010) | na | 2013^15^ | 2010/2013 | 226 | Obese mothers and healthy infants born at term (>258 days of gestation), postnatal age > 48h. Women who intended to breastfeed and had participated in the Treatment of Obese Pregnant study (TOP-study) | Breastfeeding, infant anthropometrics at 6 months of age |
| France  (Parat)^16^ | NCT00804765  (2008) | na | 2019^16^ | 2008/2013 | 275 | Women at ≤21 gestational weeks with BMI >25 kg/m^2^ | Excessive infancy weight gain defined as >0.67 change in weight SD score |
| Guatemala  (Acero)^17^ | NCT03399617  (2018) | 2020^17^ | na | 2018/2021 | 1500 | Women in third trimester or with children up to 3 months of age | Infant and young child feeding practices, height, weight gain rate, haemoglobin, obesity, stunting, anaemia |
| India  (Kumaran)^18^ | na | na | na | 2020/na | na | Women of reproductive age | Adiposity at 5 years of age |
| Ireland, UK, Spain, Australia  (O’Reilly) | ACTRN 12620001240932 (2020) | na | na | 2021/2024 | 800 | Pregnant women aged 18-50 years at high risk of developing gestational diabetes | Maternal: BMI at 12 months postpartum; Infants: weight, height, head circumference at age 12 months |
| Italy  (Maffeis)^19^ | NCT03131284  (2017) | na | 2019^19^ | 2014/2017 | 562 | Healthy full-term newborns | BMI at two years of age |
| Netherlands  (Corpeleijn)^20^ | NCT01127412  (2010) | na | 2014^20^ | 2006/2010 | 161 | Infants aged up to 2 weeks | Growth (weight, length), body composition (waist circumference, hip circumference, skinfold thickness, bioelectrical impedance analyses) all assessed regularly during the first 2.5 years of life |
| Netherlands  (Karssen)^21^ | NL6727/NTR6938  (2017) | na | na | 2018/2019 | 300 | Parents with a child aged 7-11 months | BMI at 6 and 12 months |
| Netherlands  (Mesman)^22^ | NCT03348176  (2017)  NL6397/NTR6572  (2017) | 2019^22^ | na | 2016/2020 | 255 | First-time mothers of healthy term infants 4 months to 3 years of age | Vegetable intake, vegetable liking, self-regulation of energy intake |
| Netherlands  (Raat)^23^ | NL1721/NTR1831  (2009) | 2013^24^ | 2017^23^ | 2008/2013 | 2102 | Newborn children and their parents | BMI and energy-balance related behaviours |
| New Zealand  (Taylor 2017a)^25^ | NCT00892983  (2009) | 2011^26^ | 2017^25^ | 2009/2017 | 802 | Women aged over 16 before 34 weeks’ gestation. Babies excluded if they are not full-term | Weight at 6, 12 and 24 months of age; BMI at 24 months of age |
| New Zealand  (Taylor 2017b)^27^ | ACTRN12612001133820  (2012) | 2015^28^ | 2017^27^ | 2012/2016 | 206 | Pregnant women aged 16 or over, before 34 weeks’ gestation. Babies excluded if they are not full-term | BMI at 12 months |
| Norway  (Helle)^29^ | ISRCTN13601567  (2016) | 2017^30^ | 2019^29^ | 2015/2021 | 718 | Parent and 5.5-month-old child | Child eating behaviour, child food intake, child mealtime routines, maternal feeding practices |
| Norway  (Øverby)^31^ | ISRCTN45864056  (2016) | na | 2017^31^ | 2012/2015 | 110 | Parents and 4-6 month old infant | Food intake when infants are 6, 15 and 24 months of age |
| Norway  (Røed)^32^ | ISRCTN92980420  (2017) | 2019^32^ | na | 2015/2022 | 298 | Children close to 12 months and one of their parents | Child diet quality and food variety, assessed at inclusion, 18, 24, and 48 months |
| South Africa  (Norris)^33^ | PACTR201903750173871 (2019) | na | na | 2019/2021 | 6000 | 18-25 year old women preconception and child | Dual-energy X-ray absorptiometry-derived fat mass index of the index-child at age 5 years |
| Spain  (López)^34^ | NCT03444415  (2018) | na | na | 2015/2018 | 414 | Pregnant women with a gestational age of 12-16 weeks | BMI at 2 years of age |
| Sweden  (Marcus)^35^ | NCT01198847  (2010) | 2011 ^35^ | na | 2010/2019 | 123 | At least one obese (BMI ≥ 30) or two overweight (BMI ≥ 25) parents with a one-year-old child | BMI at 6 years of age |
| Sweden  (Rasmussen)^36^ | ISRCTN16991919  (2013) | 2014^37^ | 2016^36^ | 2008/2015 | 1039 | First-time mothers | BMI and waist circumference of children & their mothers at age 4 |
| UK  (Bryant)^38^ | ISRCTN56735429  (2013) | na | 2016^38^ | 2012/2012 | 120 | Overweight/obese pregnant women (BMI ≥25) at 10–12 weeks’ gestation and infants from birth | Child’s weight |
| UK  (Bryant)^39^ | NCT03333733  (2017) | 2018^39^ | na | 2017/2019 | 115 | Parents and at least 1 child aged 6 months - 5 years | Feasibility, child BMI z-score |
| UK  (Lakshman)^40^ | ISRCTN20814693  (2011) | 2015^41^ | 2018^40^ | 2011/2015 | 669 | Parents (mainly mothers)  and their babies (aged 2 to 14 weeks) who are formula-fed | The change in infant weight standard deviation score from birth to age 12 months |
| USA  (Barlow)^42^ | NCT03101943  (2017) | na | 2020^42^ | 2017/2019 | 134 | Mothers aged ≥ 13 years, their infant aged < 14 weeks, self-reported Native American | Infant consumption of sugar-sweetened beverages, change in complementary feeding and responsive parenting practices |
| USA  (Barlow)^43^ | NCT03334266  (2017) | 2019^43^ | na | 2017/2021 | 338 | Expectant Native American mothers aged 14-24 who are having their first or second baby | Percentage of parents who meet breastfeeding and complementary feeding recommendations, child feeding styles, fruit & vegetable intake, physical activity levels, screen time, BMI |
| USA  (Beck)^44^ | NCT03438721  (2018) | na | na | 2018/2021 | 240 | Parents who self-identify as Latino and their newborn infants | Child dietary intake and screen time, parent health-related quality of life |
| USA  (Birch/Paul)^45^ | NCT00359242  (2006) | na | 2011^45^ | 2006/2009 | 160 | Mother-newborn dyads, primiparous, singleton, gestational age ≥ 34 weeks | Weight-for-length percentile at age 1 year |
| USA  (Bonuck)^46^ | NCT00756626  (2008) | na | 2014^46^ | 2008/2011 | 300 | Children 11-13 months old consuming >2 bottles of milk or juice per day | Bottle use frequency |
| USA  (Burgess)^47^ | NCT04042467  (2019) | na | na | 2019/2023 | 900 | Caregiver/infant dyads with an infant up to 14 days old | Child weight for length trajectory up to 24 months |
| USA  (Caballero)^48^ | na | na | 2015^48^ | na/na | 292 | Healthy newborns with ≥ 2000g body weight | Anthropometry (weight, height, triceps, and subscapular skinfolds), child feeding practices and dietary assessment |
| USA (Puerto Rico)  (Campos)^49^ | NCT03517891  (2018) | na | na | 2018/2022 | 480 | Low-income women in third trimester, intend to participate in the Special Nutrition Women, Infants & Children Program; infants included from birth | Adequate weight gain during the first year (gender adjusted Z score). |
| USA  (Clouiter)^50^ | NCT02052518  (2014) | 2015^51^ | 2018^50^ | 2013/2016 | 57 | Mother/newborn dyads in low-income neighbourhoods | Breastfeeding, Infant weight-for-length, BMI, introduction of solids and juice/sugar-sweetened  beverages |
| USA  (de la Haye & Salvy) ^52^ | NCT03529695  (2018) | 2019 ^52^ | na | 2018/2022 | 300 | Mother-child dyads enrolled in home visitation programs; Infants aged between birth to 24 months | Weight of mothers, rate of weight gain of infants, waist circumference of mother |
| USA  (Fiks)^53^ | NCT02037490  (2014) | na | 2017^53^ | 2014/2015 | 87 | Pregnant women, Medicaid insured, BMI ≥ 25 | Feasibility |
| USA  (Goran) ^54^ | NCT03141346  (2017) | na | na | 2017/2022 | 240 | Latina mothers who are habitual consumers of sugar sweetened beverages/juices and singleton infants less than one month | infant length, maternal height, maternal weight, maternal BMI, infant weight, infant weight z-scores, infant body composition (body fat, lean mass, total body water, free body water) at 6, 12 and 24 months |
| USA  (Groner)^55^ | NCT01565525  (2012) | 2009^56^ | 2012^55^ | 2005/2007 | 292 | Mother-infant dyads were enrolled at their first well-child visit to 3 urban paediatric clinics | Infant weight for height |
| USA  (Horodynski)^57^ | ACTRN12610000415000  (2010) | 2011^58^ | 2017^57^ | 2010/2013 | 547 | Economically and educationally disadvantaged African American, Hispanic, and Caucasian mothers with infant ≤4 months of age | Maternal responsiveness, maternal feeding style, maternal feeding practices, infant growth pattern at 6 and 12 months of age |
| USA  (Horodynski)^59^ | NCT02244424  (2014) | 2015^59^ | na | 2014/2016 | 164 | Low-income adolescent (aged 14-19 years), first-time mothers of infants ≤2 months of age | Maternal responsiveness, maternal feeding style, maternal  feeding practices, infant weight |
| USA  (Jacobvitz)^60^ | NCT04177472  (2019) | na | na | 2019/2023 | 150 | Mothers and assisting caregivers with babies 4-5 months of age | BMI percentile at 12 months of age |
| USA (Puerto Rico)  (Joshipura)^61^ | NCT01771133  (2013) | na | 2019^61^ | 2013/2015 | 31 | Overweight/obese women with a singleton pregnancy <16 weeks’ gestation and their infant from birth | Gestational weight gain |
| USA  (Karanja)^62^ | NCT00245180  (2015) | 201 ^62^ | na | 2006/2012 | 577 | American Indian infants aged 0–2 years and their mothers | BMI z-score at 24 months of age |
| USA  (Karasz)^63^ | NCT03077425  (2017) | 2018^63^ | na | 2017/2021 | 360 | Mother-child dyads where child is <6 months old | Number and amount of sippy cups and bottles per day consumed by child |
| USA  (Lavner)^64^ | NCT03505203  (2018) | 2019^64^ | na | 2018/2021 | 300 | First-time African American mothers and their full-term infants | Change in infants' weight for age from 3 weeks to 16 weeks |
| USA  (Linares)^65^ | NCT03903146 | na | 2019^65^ | 2016/2018 | 39 | Hispanic pregnant women who intended to breastfeed and their baby | Exclusive breastfeeding from birth to 6 months old |
| USA  (Messito)^66^ | NCT01541761  (2012) | na | 2016^66^ | 2012/2020 | 533 | Pregnant women with a singleton uncomplicated pregnancy | Infant feeding practices and material infant feeding knowledge |
| USA (Puerto Rico, Hawaii)  (Palacios)^67^ | NCT02903186  (2016) | na | 2018^67^ | 2016/2016 | 202 | Caregivers of healthy term infants 0-2 months participating in the Women, Infants and Children Program | Infant weight-for-length percentile |
| USA  (Paul)^68^ | NCT00125580  (2005) | na | na | 2005/2006 | 40 | Infant at ≥37 weeks' gestation,  primiparous mother, singleton, breast or bottle-fed, birth weight ≥2500g | The rate of sleeping through the night at 8 weeks of age |
| USA  (Paul)^69^ | NCT01167270  (2010) | 2014^69^ | na | 2012/2023 | 316 | Full term singleton infants born to primiparous mothers | BMI z-score at age 3 years |
| USA  (Reifsnider)^70^ | NCT01905072  (2013) | 2013^71^ | 2018^70^ | 2012/2017 | 177 | Mexican American mother, pre-pregnant BMI ≥25, aged 18-40 years; Singleton infants, >38 weeks’ gestational age, birthweight > 2500g | Child weight-for-length BMI at 3 years |
| USA  (Rothman)^72^ | NCT01040897  (2009) | na | 2014^72^ | 2010/2014 | 865 | Child presenting for 2 month well-child check-up (between 6-16 weeks of age) with an intervention-trained resident physician, caregiver able to speak Spanish or English | Percent of children overweight or obese at 2 years |
| USA  (Dutton & Salvy)^73^ | NCT03433456  (2018) | 2018^73^ | na | 2018/2022 | 596 | Mother or caregiver and 0-4 year old child | Change in mother and infants’ body weight from baseline to follow-up at 12 and 24 months |
| USA  (Savage)^74^ | NCT03482908  (2018) | 2018^74^ | na | 2016/2019 | 289 | Mother and infant <2 months, birthweight ≥ 2500g and gestational age ≥ 37 weeks | Sex-specific z-scores for infant growth measures at birth, and at 2, 5 and 7 months after birth; Sex-specific z-scores for infant rapid weight gain from birth to 6 months |
| USA  (Stephens)^75^ | NCT03249324  (2017) | 2015^75^ | na | 2014/2017 | 58 | Mothers 18-35 years of age who are eligible for care within the Military Health System; Infants from birth | Maternal weight gain |
| USA  (Stough)^76^ | NCT03597061  (2018) | na | na | 2018/2020 | 34 | Parent and infant born > 38 weeks’ gestation, above 10^th^ percentile of length-for-weight, aged 2-3 months | Weight-for-Length percentile, appetite regulation, fruit and vegetable variety at 3 and 9 months of age |
| USA  (Thomas)^77^ | NCT03601299  (2018) | na | na | 2018/2022 | 804 | Children 0-5 years of age | BMI, traditional food content |
| USA  (Thomson)^78^ | NCT01746394  (2012) | 2014^79^ | 2018^78^ | 2013/2016 | 82 | Pregnant women at least 18 years of age, < 19 weeks pregnant and their infant from birth | Maternal: weight gain at 9 months’ gestation, weight retention at 12 months postpartum, dietary intake, physical activity; Infant: dietary intake, activity, BMI at age 12 months |
| USA  (Virudachalam)^80^ | NCT01710423  (2012) | na | na | 2012/2013 | 47 | Families with children ages 0 to 3 | Change in healthfulness of the diet |
| USA  (Wasser)^81^ | NCT01938118  (2013) | 2017^82^ | 2020^81^ | 2013/2017 | 430 | Non-Hispanic black mothers enrolled at 28 weeks’ pregnancy and infants from birth to 15 months postpartum | Infants’ mean weight-for-length z-score at 15 months of age |

na = not available, BMI = body mass index
